# Testing for SARS-CoV-2 among cruise ship travelers repatriated to the United States, February–March 2020

**DOI:** 10.1101/2021.06.07.21258318

**Authors:** Michelle A. Waltenburg, Mary A. Pomeroy, Laura Hughes, Jeremy A.W. Gold, Oren Mayer, Arnold Vang, Benjamin D. Hallowell, Loretta Foster, Kerui Xu, Rita Espinoza, Kristina Hsieh, Emily G. Pieracci, Gabriella Wuyke, Juliana Da Silva, R. Paul McClung, Jonathan Steinberg, Matthew Westercamp, Snigdha Vallabhaneni, Jessica Li, Amy L. Valderrama, George R. Grimes, R. Reid Harvey, Randall J. Nett, Kindra Stokes, Stephen Lindstrom, Allison D. Miller, Eric P. Griggs, Jennifer L. Milucky, Adam Bjork, Valerie Albrecht, Wendi L. Kuhnert, Carolyn V. Gould, Nancy W. Knight, Noele P. Nelson, Margaret A. Honein, Casey Barton Behravesh, CDC COVID-19 Investigation Team, Christine L. Dubray, Grace E. Marx

**Author notes:** **Corresponding author**: Michelle Waltenburg, DVM, MPH; Centers for Disease Control and Prevention, 1600 Clifton Rd NE, Mailstop H24-10, Atlanta, GA 30329, USA. Rhode Island Department of Health, Providence, RI, USA.

## Abstract

**Background:** In early 2020, an outbreak of coronavirus disease 2019 occurred among passengers and crew of the *Diamond Princess* cruise ship. During February 16–17, some US citizens, residents, and their partners voluntarily repatriated to the US from Japan.

**Methods:** We conducted a retrospective, longitudinal evaluation of repatriated travelers where the outcome of interest was a positive test for SARS-CoV-2. Travelers who tested positive for SARS-CoV-2 were isolated in hospitals or at home under county isolation orders and underwent serial testing by real-time reverse transcription polymerase chain reaction (RT-PCR) approximately every other day, as contemporaneous US guidance required two consecutive negative tests collected ≥24 hours apart and symptom improvement before release from isolation.

**Results:** Among quarantined repatriated travelers, 14% tested positive for SARS-CoV-2. One-fifth of infected travelers initially tested negative but were identified on subsequent testing. All infected travelers remained asymptomatic or developed mild symptoms during isolation. Many travelers remained in prolonged isolation because of persistent viral detection based on contemporaneous policies.

**Conclusion:** Our findings support testing within 3-5 days after possible SARS-CoV-2 exposure to comprehensively identify infections and mitigate transmission and lend support to symptom- and time-based isolation recommendations, rather than test-based criteria.

## 1. Introduction

In early 2020, an outbreak of coronavirus disease 2019 (COVID-19) occurred among passengers and crew of the *Diamond Princess* cruise ship [1,2]. During February 16–17, 329 US citizens, residents, and their partners (subsequently referred to as repatriated travelers) voluntarily repatriated to the US from Japan. Of those repatriated, 328 were quarantined under federal authority at US military bases; one traveler was not quarantined based on documentation of having recovered from COVID-19 in Japan [1,2]. Travelers who tested positive for SARS-CoV-2, the virus that causes COVID-19, were isolated in hospitals or at home under county isolation orders and underwent serial testing by real-time reverse transcription polymerase chain reaction (RT-PCR) approximately every other day, as contemporaneous US guidance required two consecutive negative tests collected ≥24 hours apart and symptom improvement before release from isolation [3]. We report the RT-PCR results and symptom status of travelers to describe viral dynamics during mild or asymptomatic COVID-19 and to inform testing strategies after SARS-CoV-2 exposure.

## 2. Materials and methods

### 2.1 Study design and data sources

We conducted a retrospective, longitudinal evaluation of repatriated travelers; the outcome of interest was a positive test for SARS-CoV-2. Specimens were collected by oropharyngeal (OP) and/or nasopharyngeal (NP) swab and tested by RT-PCR for SARS-CoV-2 [4]. Available RT-PCR results were obtained from the Japanese Ministry of Health, Labour, and Welfare, the Centers for Disease Control and Prevention (CDC), and state public health laboratories. This activity was reviewed by CDC and was conducted consistent with applicable federal law and CDC policy.^§^

### 2.2 Demographic and symptom information

We reviewed demographic and symptom data collected at the time of the first positive SARS-CoV-2 test. Demographic characteristics for travelers who tested positive for SARS-CoV-2 were compared with travelers who tested negative; proportions for sex were compared using χ^2^ test and age compared using the Mann-Whitney *U* test method. Travelers were categorized as asymptomatic, presymptomatic (no symptoms at time of first positive result but later developed symptoms), or symptomatic (at least one new or worsened symptom of COVID-19 at time of first positive result) [5]. Mild illness was defined as individuals with any sign or symptom of COVID-19 without shortness of breath, dyspnea, or abnormal chest imaging [6].

### 2.3 Laboratory results

We calculated time to RT-PCR negativity (i.e., reversion to negative) as the number of days between the first positive RT-PCR test and the first of two consecutive negative tests on paired NP and OP specimens collected ≥24 hours apart. Intermittent viral shedding was defined as a positive RT-PCR test following a negative test. Cycle threshold (Ct) values for amplification of two targets (N1 and N2) of the viral nucleocapsid gene were analyzed for NP and OP specimens submitted to CDC (*4*). Specimens with Ct values <40 for both targets were considered positive for SARS-CoV-2 (*4*). Ct values for the N1 and N2 genetic markers were highly correlated (Appendix A); for simplicity, we reported Ct values for the N1 genetic marker.

## 3. Results

### 3.1 Study population

Of 328 quarantined repatriated travelers, 45 (14%) tested positive for SARS-CoV-2; all remained asymptomatic or developed mild symptoms during isolation. Among 45 travelers, 28 (62%) had a positive RT-PCR result in Japan before repatriation, 10 (22%) had negative results in Japan but then had a positive result in the US, and 7 (16%) who were never tested in Japan had a positive result in the US. The median time to test positivity after repatriation among the 10 travelers who tested negative in Japan was 3 days (range: 0–9). Demographic characteristics of repatriated travelers, regardless of test result, were similar (Table 1). Overall, 25 (56%) of 45 travelers were asymptomatic or presymptomatic at the time of first positive SARS-CoV-2 result.

**Table 1.**
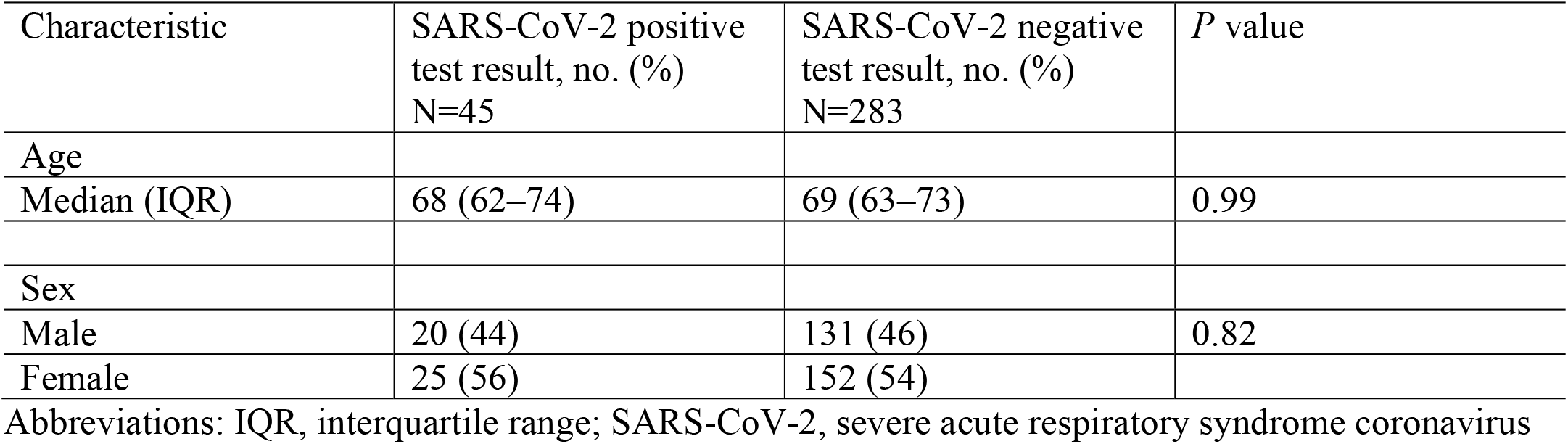
Demographic characteristics of 328 US-repatriated cruise ship travelers under federal quarantine or isolation, February to March 2020.

### 3.2 Laboratory results

Results from 556 RT-PCR tests from 45 travelers who tested positive for SARS-CoV-2 were available for analysis. Median time to RT-PCR negativity for 30 travelers for whom two consecutive negative results were available was 17 days (range = 3–38). The proportion of these 30 travelers with reversion to negative at days 10, 14, and 21 after diagnosis was 30%, 43%, and 63%, respectively (Figure 1). Two consecutive negative results were not available for 15 travelers because of missing data after travelers were issued state or county isolation orders, or released from isolation according to updated US guidance that no longer required two consecutive negative results (*3*); these 15 travelers remained in isolation ≥21 days. Intermittent viral shedding was observed in 16 (35.6%) of 45 travelers. Ct values, available from 230 SARS-CoV-2 positive RT-PCR tests from 43 travelers, increased over time after diagnosis (Figure 2).

**Figure 1.**
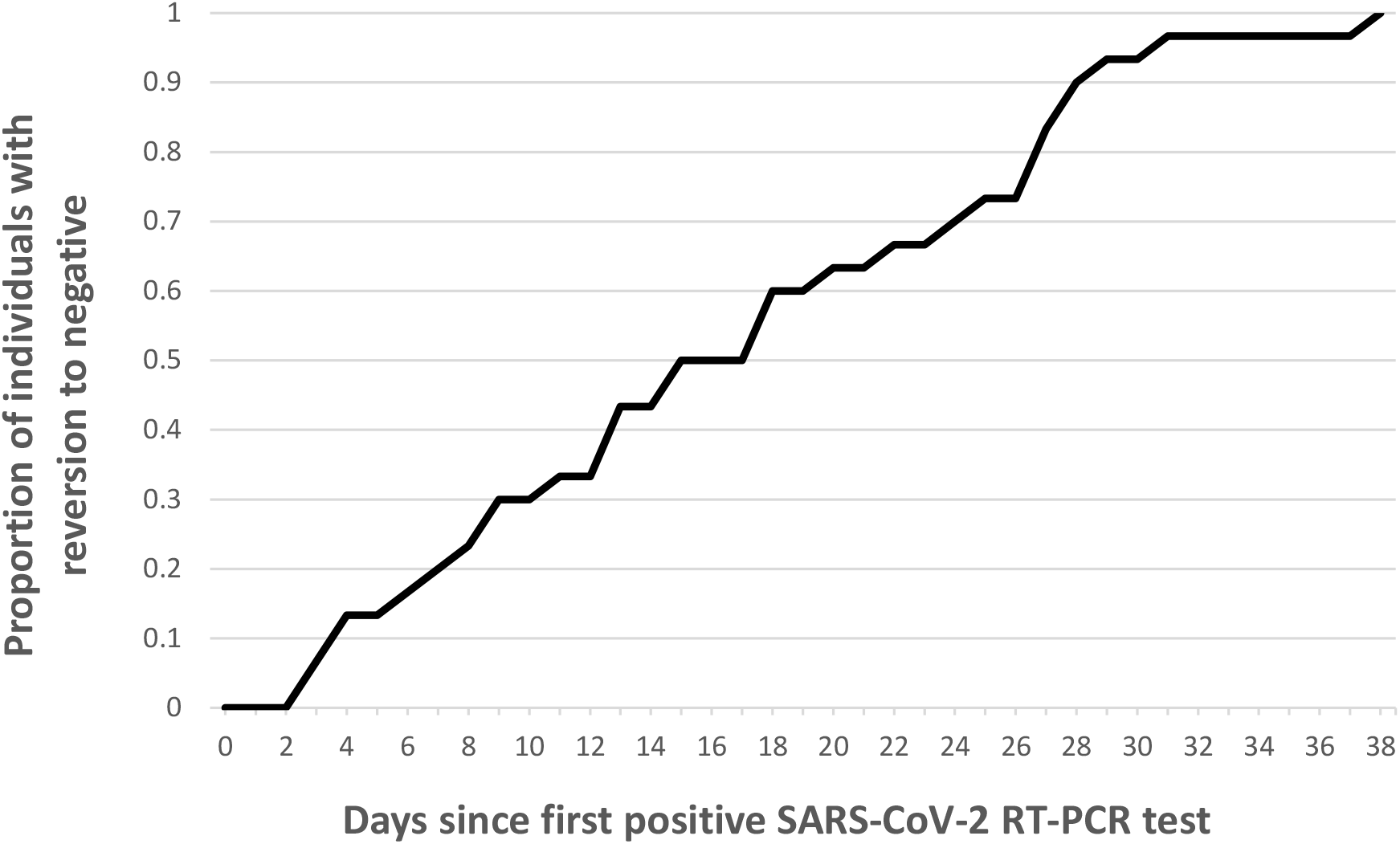
Time to reversion to negative^a^ relative to first positive SARS-CoV-2 test among 30 US-repatriated cruise ship travelers, February to March 2020. Abbreviations: SARS-CoV-2, severe acute respiratory syndrome coronavirus 2; RT-PCR, real-time reverse transcription polymerase chain reaction. ^a^Time to reversion to negative was defined as the number of days between the first SARS-CoV-2 positive RT-PCR test and the first of two consecutive negative RT-PCR tests on paired nasopharyngeal and oropharyngeal specimens collected at least 24 hours apart.

**Figure 2.**
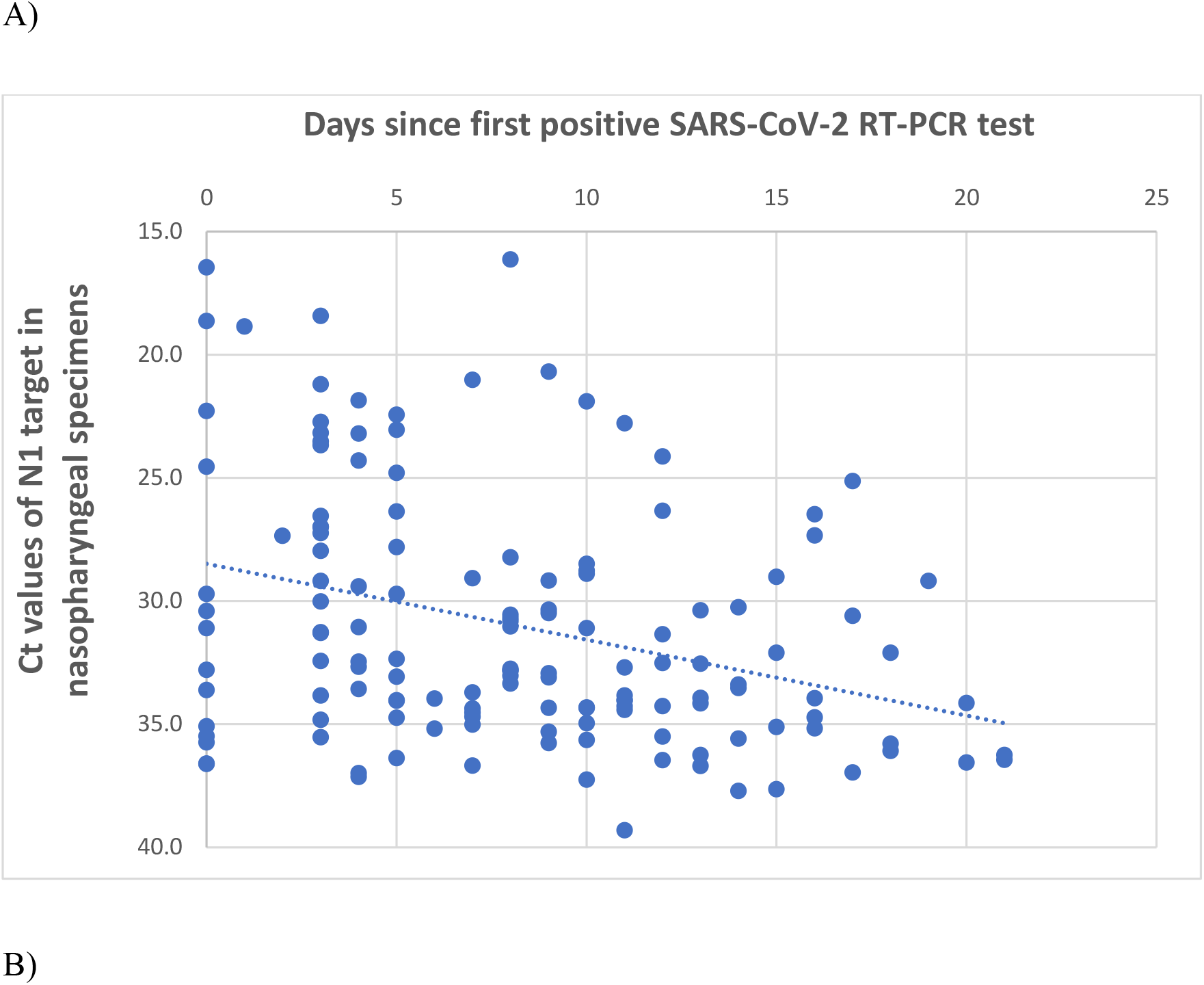
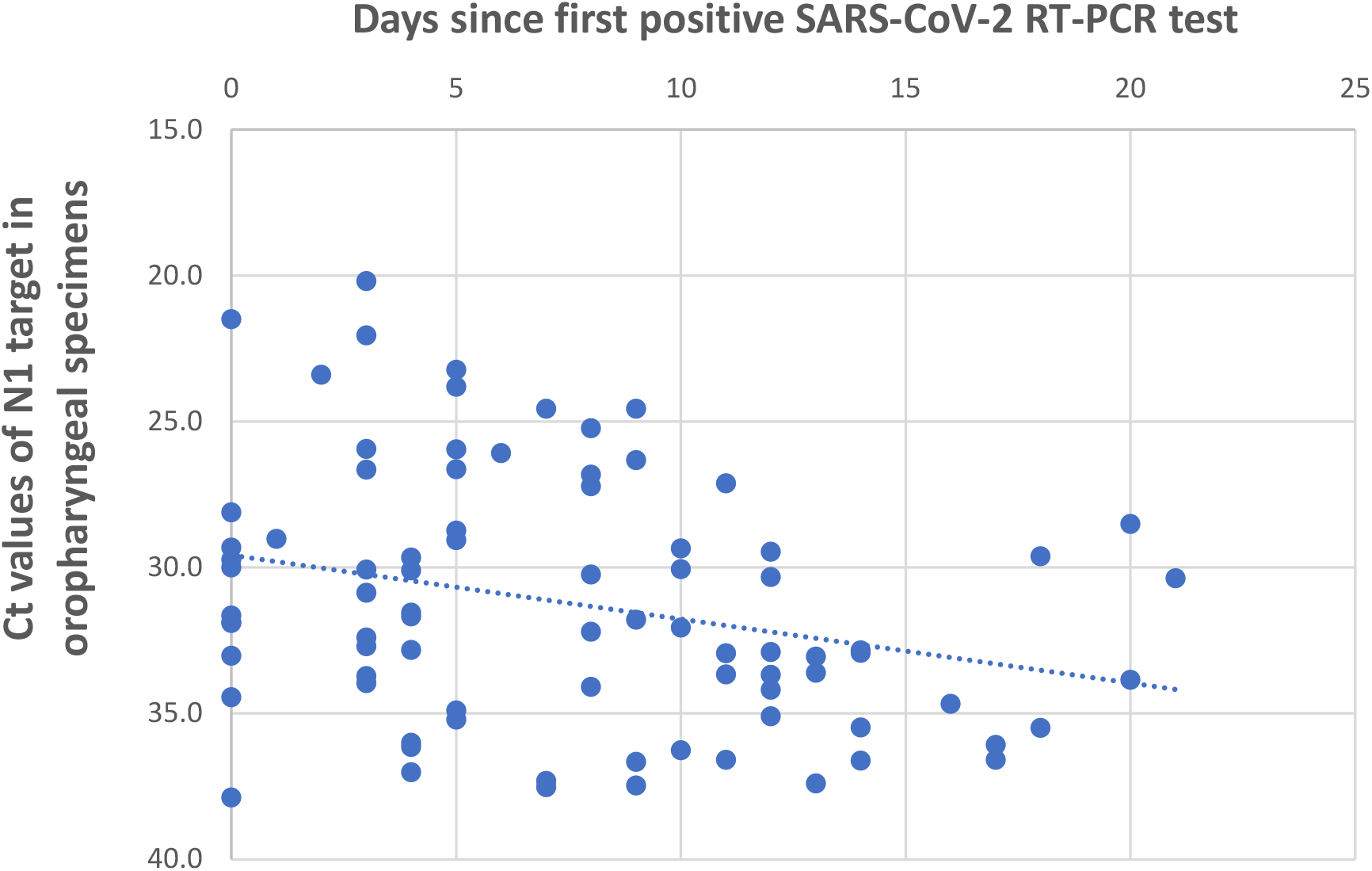
Nucleocapsid (N)1 target cycle threshold (Ct) values of SARS-CoV-2 relative to first positive test^a^ for 43 US-repatriated cruise ship travelers, February to March 2020. (A) N1 target Ct values by nasopharyngeal swab specimens (n=143). (B) N1 target Ct values by oropharyngeal swab specimens (n=83). Values <40 cycles were considered positive for SARS-CoV-2. Dotted lines represent linear regressions of N1 target Ct value versus days since first positive SARS-CoV-2 RT-PCR test. Abbreviations: SARS-CoV-2, severe acute respiratory syndrome coronavirus 2; RT-PCR, real-time reverse transcription polymerase chain reaction. ^a^Day 0 = date of first positive test.

## 4. Discussion

This report describes SARS-CoV-2 testing among 45 travelers with mild or asymptomatic infection repatriated from a single cruise ship during February–March 2020. At the time of their first positive test, half of the travelers were asymptomatic or presymptomatic. Three weeks after diagnosis, 58% remained in isolation because of prolonged viral shedding.

Our findings support testing within 3-5 days after possible SARS-CoV-2 exposure to comprehensively identify infections and mitigate transmission [7]. Repeat testing in the US identified ten COVID-19 cases among repatriated travelers who had previously tested negative in Japan; most were asymptomatic or presymptomatic at the time of US testing, highlighting that symptom-based testing after possible SARS-CoV-2 exposure is not sufficient to identify all infections to prevent further transmission [8,9]. During COVID-19 outbreaks in settings where rapid spread of SARS-CoV-2 is possible, testing, irrespective of symptoms, should be considered to identify asymptomatic and presymptomatic infections [10,11]. Universal adoption of risk mitigation strategies such as masks, hand hygiene, and physical distancing is important to prevent transmission from people who are infected but asymptomatic [9,12,13].

Most repatriated travelers in our analysis remained in isolation ≥21 days because of persistent viral detection based on contemporaneous policies [3]. Applying current symptom-and time-based guidance, these travelers would have been released from isolation 10 days after symptom onset or their first positive test; it is notable that only 30% of travelers had reverted to negative at 10 days [14]. Qualitative RT-PR tests do not measure viral load, although there is a likely inverse correlation between Ct value and the amount of viral genetic material present in the specimen [15]. The Ct value dynamics in this report substantiate prior findings that concentrations of SARS-CoV-2 RNA in upper respiratory specimens decline after initial diagnosis [12,16], as does the likelihood of recovering replication-competent virus from people who persistently shed SARS-CoV-2 RNA [16,17].

### Limitations of the study

Our analysis has several limitations. Our results are not likely to be representative of the general US population; our small sample size of repatriated travelers was generally older and did not include travelers with severe COVID-19 who were hospitalized in Japan. Comprehensive longitudinal symptom and laboratory data were not available for all travelers throughout isolation. Information on patient-related factors (e.g., underlying health conditions) that might have influenced persistent viral shedding or low Ct values was not available. Ct values can be influenced by test-related factors, including specimen quality, reaction conditions, and amplification efficiency [15]. Viral culture data are needed to demonstrate the presence of replication-competent virus to determine the duration of shedding of viable virus and implications for risk of transmission.

## 5. Conclusion

Our findings illustrate the importance of timely testing after possible SARS-CoV-2 exposure and lend support to symptom- and time-based isolation recommendations, rather than test-based criteria [14].

## Data Availability

All data referred to in the manuscript are provided.

## Acknowledgements

We thank the San Antonio Metropolitan Health District, California Department of Public Health, Solano County Department of Public Health, and other county health departments for collecting and collating epidemiologic and laboratory data; healthcare facilities and physicians for providing care for repatriated travelers; and Travis Air Force Base, Joint Base San Antonio-Lackland, University of Nebraska Medical Center, the U.S. Marshals Service, and the CDC Cruise Ship Working Group for coordinating repatriation efforts. We also thank the repatriated cruise ship travelers.

## Funding

The authors received no financial support for the research, authorship, and/or publication of this article.

## Disclaimer

The findings and conclusions in this report are those of the authors and do not necessarily represent the views of the Centers for Disease Control and Prevention.

## Declaration of Competing Interest

The authors declare no potential conflicts of interest with respect to the analysis, authorship, and/or publication of this article.

## Footnotes

^§^See e.g., 45 C.F.R. part 46, 21 C.F.R. part 56; 42 U.S.C. §241(d); 5 U.S.C. §552a; 44 U.S.C. §3501 et seq.

## Abbreviations

COVID-19: coronavirus disease 2019
SARS-CoV-2: severe acute respiratory syndrome coronavirus 2
RT-PCR: real-time reverse transcription polymerase chain reaction

---

**Supplementary Figure.**
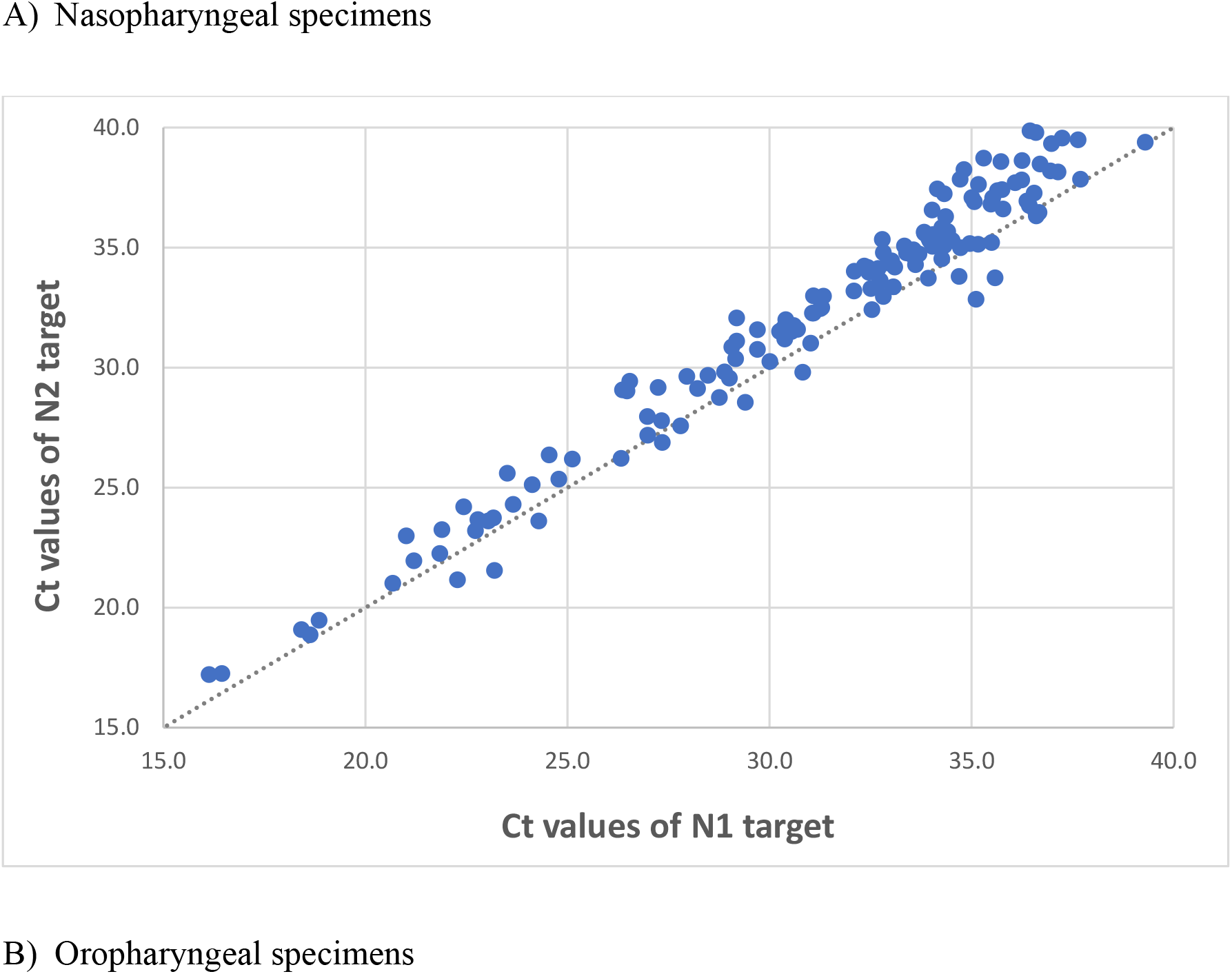
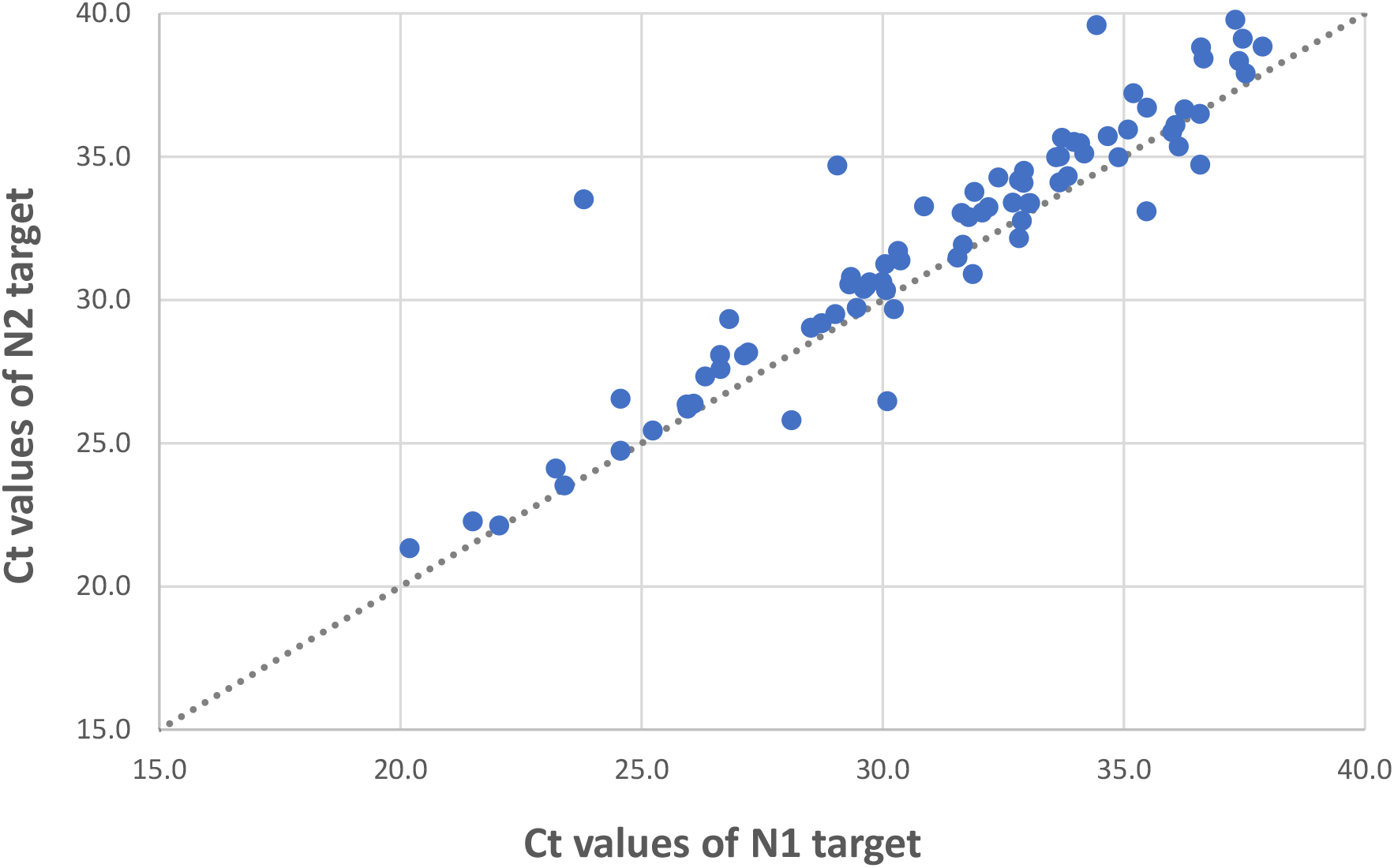
Correlation of SARS-CoV-2 nucleocapsid (N)1 and N2 target cycle threshold (Ct) values for 43 US-repatriated cruise ship travelers, February to March 2020. Each point represents the Ct values of a single nasopharyngeal (A) or oropharyngeal (B) specimen, as measured by the N1 target (x-axis) and N2 target (y-axis). For reference, perfect correspondence (where N1 and N2 target Ct values are equal) is illustrated with a dotted line. Abbreviations: SARS-CoV-2, severe acute respiratory syndrome coronavirus 2; RT-PCR, real-time reverse transcription polymerase chain reaction.

## Notes

### Competing Interest Statement

The authors have declared no competing interest.

### Author Declarations

Case investigation, data collection, and analysis were conducted for public health purposes. This project was reviewed by the National Center for Emerging and Zoonotic Infectious Diseases Human Subjects Contact at the Centers for Disease Control and Prevention (CDC). The project was determined to meet the requirements of public health surveillance covered by the U.S. Department of Health and Human Services Policy for the Protection of Human Research Subjects as defined in 45 CFR 46.102, and the decision was made that this project was nonresearch and did not require ethical review by the CDC Human Research Protection Office. Ethical approval was waived and informed consent was not required.

## References

[1] Plucinski MM, Wallace M, Uehara A, Kurbatova EV, Tobolowsky FA, Schneider ZD, et al. COVID-19 in Americans aboard the Diamond Princess cruise ship. Clin Infect Dis. 2020;ciaa1180. https://doi.org/10.1093/cid/ciaa1180

[2] Moriarty LF, Plucinski MM, Marston BJ, Kurbatova, EV, Knust B, Murray EL, et al. Public Health Reponses to COVID-19 Outbreaks on Cruise Ships – Worldwide, February–March 2020. MMWR Morb Mortal Wkly Rep 2020;69:347–52. https://doi.org/10.15585/mmwr.mm6912e3

[3] Centers for Disease Control and Prevention. Discontinuation of Isolation for Persons with COVID-19 Not in Healthcare Settings, https://www.cdc.gov/coronavirus/2019-ncov/hcp/disposition-in-home-patients.html; 2020 Mar 16 [accessed 5 Aug 2020].

[4] Centers for Disease Control and Prevention. CDC 2019-novel coronavirus (2019-nCoV) real-time RT-PCR diagnostic panel, https://www.fda.gov/media/134922/download; 2020 [accessed-15 Aug 2020].

[5] Centers for Disease Control and Prevention. Symptoms of Coronavirus, https://www.cdc.gov/coronavirus/2019-ncov/symptoms-testing/symptoms.html; 2020 Dec 22 [accessed 13 Jan 2021].

[6] Centers for Disease Control and Prevention. Discontinuation of Transmission-Based Precautions and Disposition of Patients with COVID-19 in Healthcare Settings (Interim Guidance), https://www.cdc.gov/coronavirus/2019-ncov/hcp/disposition-hospitalized-patients.html;. 2020 Aug 10 [accessed 13 Jan 2021].

[7] Centers for Disease Control and Prevention. COVID-19 and Cruise Ship Travel, https://www.nc.cdc.gov/travel/notices/covid-4/coronavirus-cruise-ship; 2020 Mar 7 [accessed 4 Mar 2021].

[8] Bagget TP, Keyes H, Sporn N, Gaeta JM. Prevalence of SARS-CoV-2 Infection in Residents of a Large Homeless Shelter in Boston. JAMA. 2020;323(21):2191–2. https://doi.org/10.1001/jama.2020.6887

[9] Moghadas SM, Fitzpatrick MC, Sah P, Pandey A, Shoukat A, Singer BH, et al. The implications of silent transmission for the control of COVID-19 outbreaks. Proc Natl Acad Sci USA. 2020;117(30):17513–5. https://doi.org/10.1073/pnas.2008373117

[10] Centers for Disease Control and Prevention. CDC Guidance for Expanded Screening Testing to Reduce Silent Spread of SARS-CoV-2, https://www.cdc.gov/coronavirus/2019-ncov/php/testing/expanded-screening-testing.html; 2021 Jan 21 [accessed 10 Mar 2021].

[11] Centers for Disease Control and Prevention. Investigating and responding to COVID-19 cases at homeless service provider sites, https://www.cdc.gov/coronavirus/2019-ncov/php/investigating-cases-homeless-shelters.html; 2020 Aug 6 [accessed 10 Mar 2021].

[12] Arons MM, Hatfield KM, Reddy SC, Kimball A, James A, Jacobs JR, et al. Presymptomatic SARS-CoV-2 infections and transmission in a skilled nursing facility. N Eng J Med. 2020;382(22):2081–90. https://doi.org/10.1056/NEJMoa2008457

[13] Centers for Disease Control and Prevention. Managing Investigations During an Outbreak, https://www.cdc.gov/coronavirus/2019-ncov/php/contact-tracing/contact-tracing-plan/outbreaks.html; 2020 Jul 31 [accessed 13 Jan 2021].

[14] Centers for Disease Control and Prevention. Discontinuation of Isolation for Persons with COVID-19 Not in Healthcare Settings, https://www.cdc.gov/coronavirus/2019-ncov/hcp/disposition-in-home-patients.html; 2021 Feb 18 [accessed 9 Mar 2021].

[15] Centers for Disease Control and Prevention. Frequently Asked Questions about Coronavirus (COVID-19) for Laboratories, https://www.cdc.gov/coronavirus/2019-ncov/lab/faqs.html; 2021 Jan 11 [accessed 13 Jan 2021].

[16] Cevik M, Tate M, Lloyd O, Maraolo AE, Schafers J, Ho A. SARS-CoV-2, SARS-CoV, and MERS-CoV viral load dynamics, duration of viral shedding, and infectiousness: a systematic review and meta-analysis. Lancet Microbe. 2021;2(1):e13–e22. https://doi.org/10.1016/S2666-5247(20)30172-5

[17] Korea Centers for Disease Control and Prevention. Findings from Investigation and Analysis of repositive cases, https://www.cdc.go.kr/board/board.es?mid=a30402000000&bid=0030&act=view&list_no=367267&nPage=1; 2020 May 19 [accessed 3 Aug 2020].

